# Brain network connectivity in major depression: extending findings from a large public data set by meta-analysis across sites

**DOI:** 10.1101/2020.06.03.20121376

**Authors:** Leonardo Tozzi, Leanne M. Williams

## Abstract

In this short communication, we test whether patients with major depression and healthy individuals have different functional connectivity within established brain networks. To this end, we leverage a very large multi-site data set of resting state fMRI data (1,300 depressed patients and 1,128 controls) collected by 25 groups. A previous study conducted on this data set compared functional connectivity of the default mode network between the two groups. In our investigation, we performed a meta-analysis across sites quantifying the effects of depression and symptom severity on connectivity of several brain-wide networks beyond the default mode. Running a meta-analysis instead of a mega-analysis also allowed us to calculate effect sizes, heterogeneity and prediction intervals that will be valuable to inform future studies wishing to investigate network functional connectivity in depression. Our results indicate that network connectivity differences between depressed and healthy subjects are consistently small, with confidence intervals almost always encompassing zero, in line with the mixed findings from previous research. Default mode network connectivity differences between depressed patients and controls were exceptionally heterogeneous across sites, suggesting the existence of depression sub-types with normo- and hypo-connected default mode network or a strong impact of clinical confounds on default mode network connectivity. The only networks for which connectivity in depressed individuals was consistently lower than in controls were the somatomotor and visual networks, which could be promising understudied targets for future investigation. Overall, we highlight the need of minimizing heterogeneity in future multi-site studies on functional connectivity in depression and the need for more research on novel taxonomies of mental illness that are robustly anchored in brain function.

## 1 Introduction

Abnormal resting-state fMRI functional connectivity within brain networks has been reported in major depressive disorder (MDD) across several studies. A previous meta-analysis (including 556 MDD and 518 controls) reported hypo-connectivity within the fronto-parietal network and hyper-connectivity within the default mode network (DMN) [1]. Recently, the Meta-MDD consortium released a very large multi-site data set of resting state fMRI data (1,300 MDD and 1,128 controls) collected by 25 groups. A previous analysis of this data set focused on the DMN and found that connectivity within it was decreased in MDD [2]. To build on these findings, we performed a meta-analysis across sites of the Meta-MDD data set quantifying the effects of MDD and symptom severity on connectivity of several canonical brain-wide networks. We calculated effect sizes, heterogeneity and prediction intervals to inform future studies.

## 2 Methods

We used the Meta-MDD participants that were not excluded after the extensive quality checks documented in [2] and excluded sites with less than 20 individuals per group. We downloaded resting state BOLD time series extracted from Power atlas [3] regions after these preprocessing steps: slice timing correction, realignment, covariates removed (with global signal regression), spatial normalization, filter (0.01-0.1Hz) ([2] for details). We censored time-points with > 0.25 frame-wise displacement and excluded subjects with > 25% of time-points censored. We computed the average Pearson correlation of time series within the networks defined by Power: default mode (DMN), salience (SAL), cingulo-opercular (COP), dorsal attention (DAN), ventral attention (VAN), fronto-parietal (FPN), memory retrieval (MEM), somato-motor hand (SMH), somato-motor mouth (SMM), visual (VIS), auditory (AUD), subcortical (SC), cerebellum (CER). We regressed out the effects of age and sex on functional connectivity for each network and site separately. Using the R package “meta”, we ran three meta-analyses for each network to pool data across sites, testing the following effects: mean connectivity of MDD < Controls (N=695 and 642), Spearman correlations between connectivity and depression severity (total Hamilton depression scale; HAMD, N=1280) [4] and anxiety (Hamilton anxiety scale; HAMA, N=995) [5]. Meta-analyses were random-effects using the Hartung-Knapp method and tau estimation using REML [6]. We corrected p-values using false discovery rate (FDR) across networks. All code is available on Github at: https://github.com/leotozzi88/meta_mdd_metaanalysis.

## 3 Results

See Table 1 for a summary of findings. Connectivity was significantly lower in MDD compared to controls in the SMM (Hedges’ g=-0.21, 95% CI=[-0.30,-0.11], p_FDR_=0.01, prediction interval=[-0.30,-0.11], I^2^=0). VIS connectivity was also decreased, but not significantly (Hedges’ g=-0.13, 95% CI=[-0.24,-0.03], p_FDR_=0.14, prediction interval=[-0.24,-0.02], I^2^=0). Sites were strongly heterogeneous regarding differences in DMN connectivity (I^2^=0.61, Figure 1) and correlations between connectivity and HAMD or HAMA (median I^2^=0.43 and 0.39 across networks).

**Table 1:** Meta-analysis results. Three meta-analyses were run for each functional network, testing the following effects: mean connectivity of MDD < Controls, Spearman correlations between connectivity and total HAMD and total HAMA. Age and sex effects were removed using regression for each site and network. Confidence and prediction intervals for effects are given in square brackets. Abbreviations: AUD=auditory, CER=cerebellum, COP=cingulo-opercular, DAN=dorsal attention, DMN=default mode, FDR=false discovery rate, FPN=fronto-parietal, HAMA=Hamilton anxiety scale, HAMD=Hamilton depression scale, g=Hedges’ g, MDD=major depressive disorder, MEM=memory retrieval, SAL=salience, SC=subcortical, SMH=somato-motor hand, SMM=somato-motor mouth, VAN=ventral attention, VIS=visual.

| Network | MDD < Controls (N=695 and 642, sites=11) |  |  |  | HAMD (N=1280, sites=11) |  |  |  | HAMA (N=995, sites=7) |  |  |  |
| --- | --- | --- | --- | --- | --- | --- | --- | --- | --- | --- | --- | --- |
| | Effect (g) | $p_{FDR}$ | Prediction | $I^2$ | Effect (g) | $p_{FDR}$ | Prediction | $I^2$ | Effect (g) | $p_{FDR}$ | Prediction | $I^2$ |
| AUD | -0.06 [-0.19,0.06] | 0.35 | [-0.31,0.19] | 0.02 | -0.02 [-0.14,0.09] | 0.90 | [-0.34,0.29] | 0.57 | 0.03 [-0.08,0.15] | 0.72 | [-0.20,0.27] | 0.40 |
| CER | -0.03 [-0.17,0.12] | 0.68 | [-0.32,0.26] | 0.18 | -0.01 [-0.10,0.07] | 0.90 | [-0.19,0.16] | 0.31 | 0.03 [-0.08,0.13] | 0.72 | [-0.20,0.25] | 0.39 |
| COP | -0.12 [-0.30,0.05] | 0.23 | [-0.48,0.23] | 0.38 | -0.06 [-0.20,0.08] | 0.74 | [-0.47,0.35] | 0.68 | 0.00 [-0.14,0.15] | 0.94 | [-0.31,0.31] | 0.55 |
| DMN | -0.19 [-0.40,0.02] | 0.17 | [-0.75,0.38] | 0.61 | -0.01 [-0.14,0.13] | 0.96 | [-0.39,0.38] | 0.68 | 0.03 [-0.10,0.15] | 0.72 | [-0.23,0.29] | 0.48 |
| DAN | -0.08 [-0.20,0.05] | 0.26 | [-0.23,0.08] | 0.00 | 0.08 [-0.07,0.24] | 0.74 | [-0.40,0.56] | 0.77 | 0.10 [0.00,0.20] | 0.72 | [-0.06,0.26] | 0.29 |
| FPN | -0.09 [-0.19,0.00] | 0.17 | [-0.19,0.01] | 0.00 | -0.03 [-0.11,0.05] | 0.74 | [-0.18,0.11] | 0.24 | 0.04 [-0.10,0.18] | 0.72 | [-0.28,0.36] | 0.55 |
| MEM | -0.03 [-0.17,0.10] | 0.62 | [-0.23,0.16] | 0.07 | -0.08 [-0.18,0.02] | 0.74 | [-0.32,0.17] | 0.47 | -0.04 [-0.26,0.18] | 0.72 | [-0.64,0.56] | 0.80 |
| SAL | -0.11 [-0.24,0.01] | 0.17 | [-0.31,0.09] | 0.00 | -0.05 [-0.13,0.03] | 0.74 | [-0.20,0.10] | 0.21 | -0.02 [-0.08,0.04] | 0.72 | [-0.08,0.04] | 0.00 |
| SMH | -0.11 [-0.23,0.02] | 0.17 | [-0.23,0.02] | 0.00 | -0.07 [-0.15,0.01] | 0.74 | [-0.21,0.07] | 0.24 | -0.05 [-0.14,0.04] | 0.72 | [-0.20,0.11] | 0.18 |
| SMM | -0.21 [-0.30,-0.11] | 0.01 | [-0.30,-0.11] | 0.00 | -0.03 [-0.12,0.07] | 0.79 | [-0.24,0.18] | 0.43 | -0.03 [-0.16,0.10] | 0.72 | [-0.32,0.26] | 0.52 |
| SC | -0.12 [-0.23,0.00] | 0.17 | [-0.24,0.00] | 0.00 | -0.04 [-0.15,0.07] | 0.79 | [-0.31,0.24] | 0.51 | -0.06 [-0.16,0.03] | 0.72 | [-0.23,0.10] | 0.20 |
| VAN | -0.06 [-0.15,0.04] | 0.26 | [-0.15,0.04] | 0.00 | 0.00 [-0.10,0.10] | 0.96 | [-0.22,0.21] | 0.44 | -0.08 [-0.22,0.07] | 0.72 | [-0.44,0.29] | 0.72 |
| VIS | -0.13 [-0.24,-0.03] | 0.14 | [-0.24,-0.02] | 0.00 | -0.04 [-0.12,0.03] | 0.74 | [-0.18,0.09] | 0.22 | -0.04 [-0.12,0.04] | 0.72 | [-0.12,0.05] | 0.04 |

**Figure 1:**
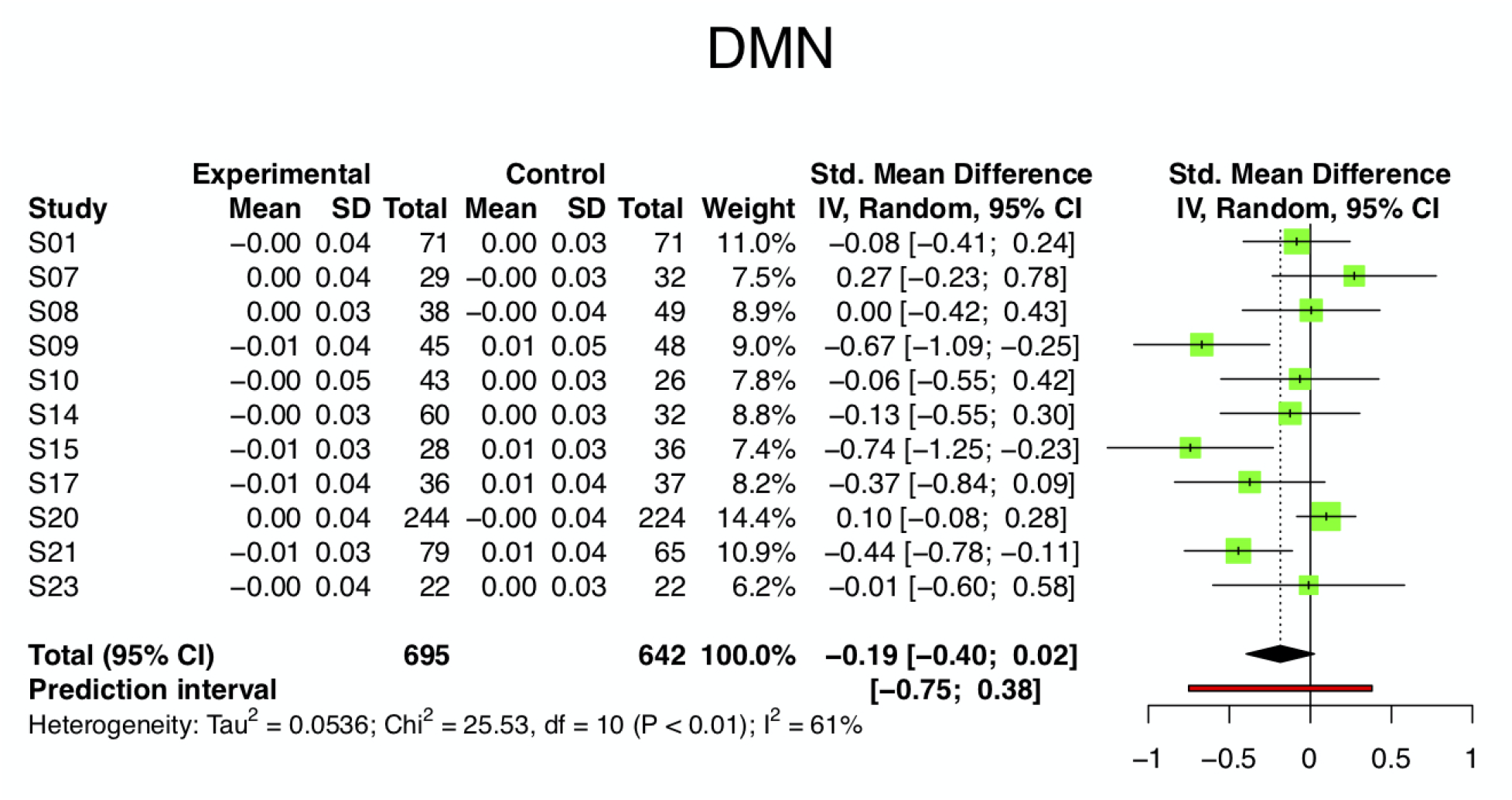
Meta-analysis result comparing average DMN connectivity between controls and MDD across sites. Age and sex effects were removed using regression for each site and network. Abbreviations: CI=confidence interval, DMN=default mode network, IV=inverse variance, SD=standard deviation.

## 4 Discussion

In this large meta-analysis across sites, we show that network connectivity differences between depressed and healthy subjects are consistently small, with confidence intervals almost always encompassing zero. This mirrors the mixed findings from previous research [1, 2]. Results were homogeneous across samples for all networks, with the exception of DMN. This might indicate the existence of MDD sub-types with normo- and hypo-connected DMN [7], or could be due to confounds like illness duration and treatment [2]. The only networks for which connectivity in MDD was lower than in controls were SMM and VIS. Importantly, prediction intervals indicate that future studies investigating average connectivity in networks other than SMM and VIS should expect no differences between MDD and controls. Concerning symptom severity, results were null and strongly heterogeneous across sites. Future multi-site studies measuring average network functional connectivity in depression should minimize heterogeneity (e.g. by harmonizing recruitment criteria and MRI sequences). Also, SMM and VIS could be promising understudied targets for future investigation. In general, more research is needed on novel taxonomies of mental illness that are robustly anchored in brain function.

## Data Availability

Data was downloaded from the publicly available Meta-MDD dataset: http://rfmri.org/REST-meta-MDD Code for the analysis is available on Github: https://github.com/leotozzi88/meta_mdd_metaanalysis

